# Increased resting-state brain entropy (BEN) in mild to moderate depression was decreased by nonpharmacological treatment

**DOI:** 10.1101/2024.04.26.24306327

**Authors:** Donghui Song, Panshi Liu, Xiaoye Ma, Da Chang, Ze Wang

## Abstract

Brain entropy (BEN) reflects the irregularity of brain activity. The neural mechanisms of depression of nonpharmacological treatments remain largely elusive. We employed BEN to evaluate neural changes in mild to moderate depression and their modulation by nonpharmacological interventions. Resting-state fMRI assessed BEN in 29 patients with mild to moderate depression at baseline and follow-up. Among them, 14 received nonpharmacological treatment, while 15 remained untreated. BEN was also evaluated in 20 matched healthy controls (HCs). Baseline BEN differences between patients and HCs were identified using two-sample t-tests. Significant regions were selected as regions of interest (ROIs). Repeated measures ANOVA tested treatment-induced BEN changes in these ROIs. The results indicate that compared to HCs, patients showed higher baseline BEN in the left dorsomedial prefrontal cortex (DMPFC), left amygdala (AMY)/putamen, and right hippocampus/parahippocampal cortex (HPC/PHPC). Treated patients had higher baseline BEN in the DMPFC than untreated patients. At follow-up, treated patients showed reduced BEN across all ROIs and reduced depression levels as measured by Beck Depression Inventory (BDI). In contrast, untreated patients demonstrated no changes and remained significantly higher BEN than HCs across all ROIs. Further analysis showed that higher BEN in DMPFC and AMY/putamen recovered to levels comparable to HCs, though BDI remained higher in treated patients. These findings suggest that bottom-up emotion dysregulation in depression is effectively mitigated by nonpharmacological treatment targeting top-down emotion regulation mediated by the prefrontal-limbic network. This study reveals that BEN can serve as an effective neural biomarker throughout the onset, progression, and recovery processes of depression.

**Highlights:** Depression is linked to increased brain entropy in prefrontal-limbic network. Nonpharmacological treatment normalizes elevated BEN of prefrontal-limbic network in depression.

Prefrontal-limbic BEN can serve as an effective neural biomarker for onset, progression, and recovery processes of depression.

## 1. Introduction

Entropy refers to the irregularities and disturbances of the dynamic system (Clausius, 1862). The second law of thermodynamics states that entropy increases over time, eventually reaching a state of maximum chaos and disorder in an isolated system. In information theory, entropy quantifies uncertainty, surprise, and the amount of information (Shannon, 1948). A living system is a highly self-organizing system that constantly interacts with the external environment to reduce its entropy to maintain proper functioning (Odum, 1988; Sneyd, Theraula, Bonabeau, Deneubourg, & Franks, 2001). The human brain, one of the most complex systems in the world, consumes a disproportional amount of body energy even without performing any overt tasks (Magistretti & Allaman, 2015), which has been postulated to maintain the brain’s normal functional state (De Ridder, Vanneste, & Freeman, 2014; K. Friston, Kilner, & Harrison, 2006) and probably maintain the balance of brain entropy (BEN) (Wang, 2021). Therefore, BEN can serve as a metric to depict the complex dynamic state of the brain associated with normal and disordered brain function.

In the human brain, regional BEN has been increasingly assessed using resting state fMRI (rs-fMRI), providing diverse information such as regional perfusion and the amplitude of low-frequency fluctuations (D. Song, Chang, Zhang, Ge, et al., 2019; D. Song, Da Chang, Ge, Zang, & Wang, 2018). Using rs-fMRI from a large cohort of healthy subjects, we previously identified normal BEN distributions at rest (Wang, Li, Childress, & Detre, 2014) and their associations with age, sex, education, and neurocognition (Wang, 2021), even after controlling for motion and physiological variables (Del Mauro & Wang, 2023). The neurocognitive correlates of resting BEN were further indicated by the correlations between regional BEN and the magnitude of task activations and task de-activations in the corresponding activated and de-activated brain regions (L. Lin, Chang, Song, Li, & Wang, 2022). Task-induced BEN, including movie-watching, also reveals the cognitive dynamics of the brain (Camargo, Del Mauro, & Wang, 2024; D.-H. Song & Wang, 2024a). The clinical and translational research suggests that BEN alterations are associated with various brain diseases (Del Mauro et al., 2025; Del Mauro, Sevel, Boissoneault, & Wang, 2024; Fu et al., 2023; Ji et al., 2022; Jiang, Cai, & Wang, 2023a, 2023b; Kuang et al., 2021; Li, Fang, Hager, Rao, & Wang, 2016; X. Liu et al., 2020; Sokunbi et al., 2013; Wang & Initiative, 2020; Xue, Yu, Guo, Song, & Wang, 2019; Zhou et al., 2014; Zhou et al., 2016), including major depressive disorder (MDD) (C. Lin et al., 2019; X. Liu et al., 2019, 2020). Importantly, we have shown that regional BEN can be modulated by repetitive transcranial magnetic stimulations (rTMS) (Chang et al., 2018; P. Liu et al., 2025; D. Song, Chang, Zhang, Peng, et al., 2019; D. Song, Deng, Chang, & Wang, 2025), caffeine (Chang et al., 2018), and medication (X. Liu et al., 2019, 2020).

Depression is a common mental disorder characterized by persistent sadness and lack of interest or pleasure in previously rewarding or enjoyable activities. Despite numerous research efforts, the brain mechanisms of this extremely inhomogeneous disease remain unclear (Chaudhury, Liu, & Han, 2015). By characterizing the temporal dynamics of the brain, BEN could open a unique window to advance our understanding of the regional neural mechanisms of depression, offering an opportunity to pinpoint region-specific targets for future intervention studies. Our recent research suggests that, in comparison to the healthy controls (HCs), the MDD group has reduced BEN in the medial orbitofrontal cortex (MOFC)/subgenual cingulate cortex (sgACC), which is a component of the default mode network (DMN) and increased BEN was observed in the motor cortex (MC). Interestingly, lower BEN in MOFC/sgACC was associated with lower levels of depression, while higher BEN in MC was associated with lower disease severity (X. Liu et al., 2019, 2020). The seemingly contradictory results between the cross-sectional comparisons and the within-subject correlations were also reported by Fu et al. (Fu et al., 2023) who observed reduced BEN in the right lateral orbitofrontal cortex (LOFC), but found that higher BEN therein was related to higher disease severity in MDD patients. In another entropy study (C. Lin et al., 2019), Lin et al. reported higher BEN in the left frontoparietal network (FPN) in late-life depressed older adults compared to HCs, which was related to lower geriatric depression scale scores (C. Lin et al., 2019). While these studies clearly demonstrated the potential of BEN for identifying regional changes and their associations with depression, the cross-sectional differences in BEN were all opposite to the within-patient BEN-disease correlation results. The within-patient BEN-disease correlation results also showed discrepancy between studies. These inconsistencies could be caused by various factors common in MDD research, including use of different antidepressants, medication history, recurrence, disease status during MRI scanning, and heterogeneity between individuals, which have been reported and discussed in other studies (Klooster & Siddiqi, 2023; Schmaal et al., 2020; Yan et al., 2019).

To avoid these often difficult-to-control confounds, we aim to focus on BEN in patients with mild to moderate depression, which can effectively avoid the influence of medication and medication history. Using a mild to moderate depression cohort may help provide clues to the early onset and progression of depression as an affective/mood spectrum (Hudson et al., 2003). We also aimed to answer a clinically important question of whether BEN can track treatment effects through nonpharmacological treatment, as we previously demonstrated that BEN can monitor the effectiveness of pharmacological treatment in MDD (X. Liu et al., 2019, 2020).

## 2. Methods and Materials

### 2.1 Dataset and demographic information of participants

We identified two datasets from OpenNeuro: ds002748 ( https://openneuro.org/datasets/ds002748) and ds003007 ( https://openneuro.org/datasets/ds003007). Both datasets were released by the same research group and acquired using identical MRI scanning parameters. We identified 29 adult patients with mild to moderate depression (DEP) (mean age = 32.83±9.06 years, 21 female), diagnosed according to the *International Classification of Diseases, Tenth Revision* (ICD-10) (Organization, 1994), from dataset ds003007. Additionally, 20 matched HCs (age = 32.90±7.40 years, 14 female) were included from dataset ds002748, excluding one individual with Raven’s scores below 70. All HCs had no history of neurological or psychiatric disorders. Fourteen depression patients (Treatment) (age = 30.29±8.36, 11 female) received nonpharmacological treatment (8 patients received brief cognitive-behavioral therapy, 6 patients underwent a real-time fMRI neurofeedback course) were scanned before and after treatment and 15 untreated depression patients (Non-Treatment) (age = 35.20±9.03, 10 female) were scanned twice with 2-3 months interval between the recordings without any treatment received. No significant differences were observed between the depression (DEP) and HCs in terms of age (Mann-Whitney *U* = 276, *p* = 0.783) or gender distribution (χ*²* = 0, *p* = 1.000). Similarly, there were no significant demographic differences (age or sex) among the Treatment, Non-Treatment, and HCs. Detailed demographic characteristics are presented in Table 1.

**Table 1.** Demographic information of participants.

| Variables | Treatment | Non-treatment | HCs | *P1* | *P2* | *P3* |
| --- | --- | --- | --- | --- | --- | --- |
| Age (M±SD) | 30.29±8.36 | 35.20±9.03 | 32.90±7.40 | 0.358 | 0.428 | 0.155 |
| Sex (f/m) | 11/3 | 10/5 | 14/6 | 0.704 | 1 | 0.682 |

Note: M ± SD: Mean ± Standard Deviation, f/m: female/male, *P1*: Treatment (n=14) vs HCs (n=20); *P2*: Non-Treatment (n=15) vs HCs (n=20). *P3*: Treatment (n=14) vs Non-Treatment (n=15).

### 2.2 Clinical Measures and Treatments

Zung Self Rating Depression Scale (SDS) (Zung, 1965) and Beck Depression Inventory (BDI) (Beck, Steer, & Brown, 1987) were used to assess depression severity. Additionally, the Rumination Response Scale (RRS) (Spasojević & Alloy, 2001) was used to assess the degree of rumination.

Cognitive-behavioral therapy (CBT): Each participant received 8 common and 8 personalized treatment sessions. Five patients were included in the common treatment sessions led by a psychiatrist and a clinical psychologist at the time. Each common session contained a course describing automatic thought, the connection between automatic thought and emotions, cognitive biases, thought modification techniques, positive reappraisal, and assertiveness. Personalized treatment sessions involved personalized and symptom-focused intervention, problems detection, work with priorities, belief-emotion connections, automatic thoughts, and cognitive biases detection and modification.

Neurofeedback (NFB): The training was designed to enhance patients’ ability to regulate left dorsomedial prefrontal cortex (DMPFC) which is supposed to be involved in positive emotions regulation through connections to amygdala (AMY). The total scanning time was approximately 30 minutes for each session. Five minutes were spent on the placement of participants into scanner and acquisition of reference images, and 25 minutes were dedicated to neurofeedback per session. In even sessions, participants spent 10 minutes of neurofeedback time on a transfer run in which they received no feedback and had to rely on their established strategies of signal regulation. In total, each participant received 8 individual sessions.

Detailed information on datasets can be obtained from previous publications (D. Bezmaternykh et al., 2018; D. D. Bezmaternykh et al., 2021; Mel’nikov et al., 2017; Ray, Bezmaternykh, Mel’nikov, Friston, & Das, 2021).

### 2.2 MRI acquisition and Preprocessing

MRI images including anatomical and functional images were acquired in the International Tomography Center, Novosibirsk, using a 3LT Ingenia scanner (Philips Inc). The anatomical images were acquired with a T1-weighted 3D turbo field echo sequence with a voxel size of 1×1×1 mm^3^. fMRI images were acquired using a T2*-weighted echo planar imaging sequence. Acquisition parameters were as follows: voxel size = 2×2×5 mm^3^, repetition time = 2.5s, echo time = 35ms. Participants were asked to lie still and motionless with eyes closed during 4 min resting state sequence and 100 volumes were required.

MRI images were preprocessed using custom scripts based on FSL (version=6.06.5) (Smith et al., 2004), Nilearn (version=0.10.2) (https://nilearn.github.io/stable/index.html) and Nipype (https://nipype.readthedocs.io/en/latest/) (K. Gorgolewski et al., 2011) applying standard preprocessing steps. First, the structural images were segmented into gray matter (GM), white matter (WM), and cerebrospinal fluid (CSF) using FAST (Zhang, Brady, & Smith, 2001), and then normalized to MNI152 space using FLIRT (Greve & Fischl, 2009; Jenkinson, Bannister, Brady, & Smith, 2002; Jenkinson & Smith, 2001). The functional images underwent preprocessing through the following steps: (1) The first two points were discarded to remove signal from the non-steady state. (2) Slice timing was corrected for the remaining time points. (3) Head motion correction was performed using MCFLIRT (Jenkinson et al., 2002), generating motion parameter files and an average functional image. (4) FLIRT was used to register the segmented structural image to the average functional image to extract signal of WM and CSF. (5) The average signals of WM and CSF were extracted from the functional image according to segmented structural brain. (6) Framewise displacement (FD) (Power, Barnes, Snyder, Schlaggar, & Petersen, 2012) was computed based motion parameter, and the mean FD was calculated using Nipype. (7) Detrending and temporal bandpass filtering (0.01-0.1 Hz) was performed. Nuisance components were regressing out including the six motion parameters, as well as the average WM signals and average CSF signals. (8) Functional images were smoothed with a 6 mm full width at half maximum (FWHM) Gaussian kernel.

### 2.3 BEN mapping calculation

The BEN mapping toolbox (BENtbx) (Wang et al., 2014) was used to calculate BEN at each voxel of the preprocessed rs-fMRI data using sample entropy (SampEn) (Richman & Moorman, 2000). The toolbox can be found at https://www.cfn.upenn.edu/zewang/BENtbx.php and https://github.com/zewangnew/BENtbx. More details of BEN calculation can be found in our original BENtbx paper or previous studies (L. Lin et al., 2022; X. Liu et al., 2020; D. Song, Chang, Zhang, Ge, et al., 2019; D. Song & Wang, 2025; Wang, 2021; Wang & Initiative, 2020; Wang et al., 2014). In this study, the window length was set to 3 and the cut off threshold was set to 0.6 based on optimized parameters (Wang et al., 2014). BEN maps were then normalized to MNI space for group analysis. To reduce variability due to noise, BEN maps were smoothed with an isotropic Gaussian kernel (FWHML=L10 mm) and resampled to 3×3×3 mm^3^.

### 2.4 Statistical analysis

#### 2.4.1 Demographic information and clinical measures analysis

Normality tests (Shapiro-Wilk) and homogeneity of variance tests (Levene’s test) were first performed. Based on these results, either independent *t*-tests or Mann-Whitney U tests were selected to examine between-group differences in age and clinical measures. Within-group changes between baseline and follow-up were assessed using paired *t*-tests. Sex was compared using chi-square tests across different groups. All statistical analyses were conducted using custom python scripts.

#### 2.4.2 BEN analysis

Whole-brain voxel-wise comparisons of baseline BEN between DEP and HCs were conducted using Statistical Parametric Mapping (SPM12; Wellcome Trust Centre for Neuroimaging, London, UK) (K. J. Friston et al., 1994), with age, sex, and mean framewise displacement (mFD) included as nuisance covariates. Statistical thresholds were set at uncorrected voxel-wise *p* < 0.001 with cluster size > 20 voxels. Regions showing significant differences between group differences were defined as regions of interest (ROIs). Mean BEN values were then extracted from each ROI for all participants. These values were submitted to 2 (Treatment: treatment vs non-treatment) × 2 (Time: baseline vs follow-up) repeated-measures analysis of variance (ANOVA) to examine treatment-by-time interactions. For ROIs demonstrating significant interactions (*p* < 0.05), then simple analyses were performed. Comparisons of BEN differences between the Treatment and Non-treatment (at both baseline and follow-up) and HCs were performed using two sample *t*-tests.

#### 2.4.3 Neurosynth decoding

To mitigate the potential statistical bias due to the small sample size, we used Neurosynth (https://neurosynth.org) to decode the functional terms associated with the unthresholded *t*-maps of BEN difference between DEP and HCs. Neurosynth (https://neurosynth.org) is a meta-analytic tool that contains 14,371 functional neuroimage studies with meta-analyses of 1,334 terms (Yarkoni, Poldrack, Nichols, Van Essen, & Wager, 2011). The analysis method can be found at https://neurosynth.org/analyses/terms/. We first uploaded unthresholded *t*-maps to NeuroVault (K. J. Gorgolewski et al., 2015) and then used the Neurosynth decoder function provided by NeuroVault to compute a voxel-wise Pearson correlation coefficient between the *t*-value and each of the term-based z-statistic maps extracted from Neurosynth. Out of the 1,334 terms, the first 100 terms are initially selected based on positive or negative correlation strength, after all anatomical, redundant, and methodologic terms were excluded, then top 30 terms were visualized as word clouds.

## 3. Results

### 3.1 Clinical measures analysis

At baseline, both the Treatment and Non-treatment groups within the DEP cohort showed significantly higher scores than HCs on the SDS, BDI, and RRS (all *p* < 0.001). Additionally, the Treatment exhibited higher scores than the Non-Treatment on the SDS (*p* = 0.027) and BDI (*p* = 0.07) but showed no significant difference in rumination as measured by the RRS (*p* = 0.150).

Following intervention, the Treatment exhibited a significant reduction in BDI scores (*p* = 0.021), while the Non-Treatment showed no significant changes at follow-up. Neither the Treatment nor Non-Treatment showed significant changes in SDS or RRS scores at follow-up. At follow-up, both groups continued to show significantly higher scores than HCs on the SDS (*p* < 0.001 for Treatment, *p* = 0.003 for Non-Treatment) and BDI (*p* < 0.001 for Treatment, *p* = 0.003 for Non-Treatment), but the Non-Treatment showed no significant difference from HCs on RRS scores (*p* = 0.102). Notably, at follow-up, there were no significant differences between the Treatment and Non-Treatment on the SDS (*p* = 0.118) or BDI (*p* = 0.658), while Treatment maintained higher rumination levels as measured by the RRS compared to the Non-Treatment (*p* = 0.024). Additional analyses of clinical measure score differences are presented in Table 2.

**Table 2.**
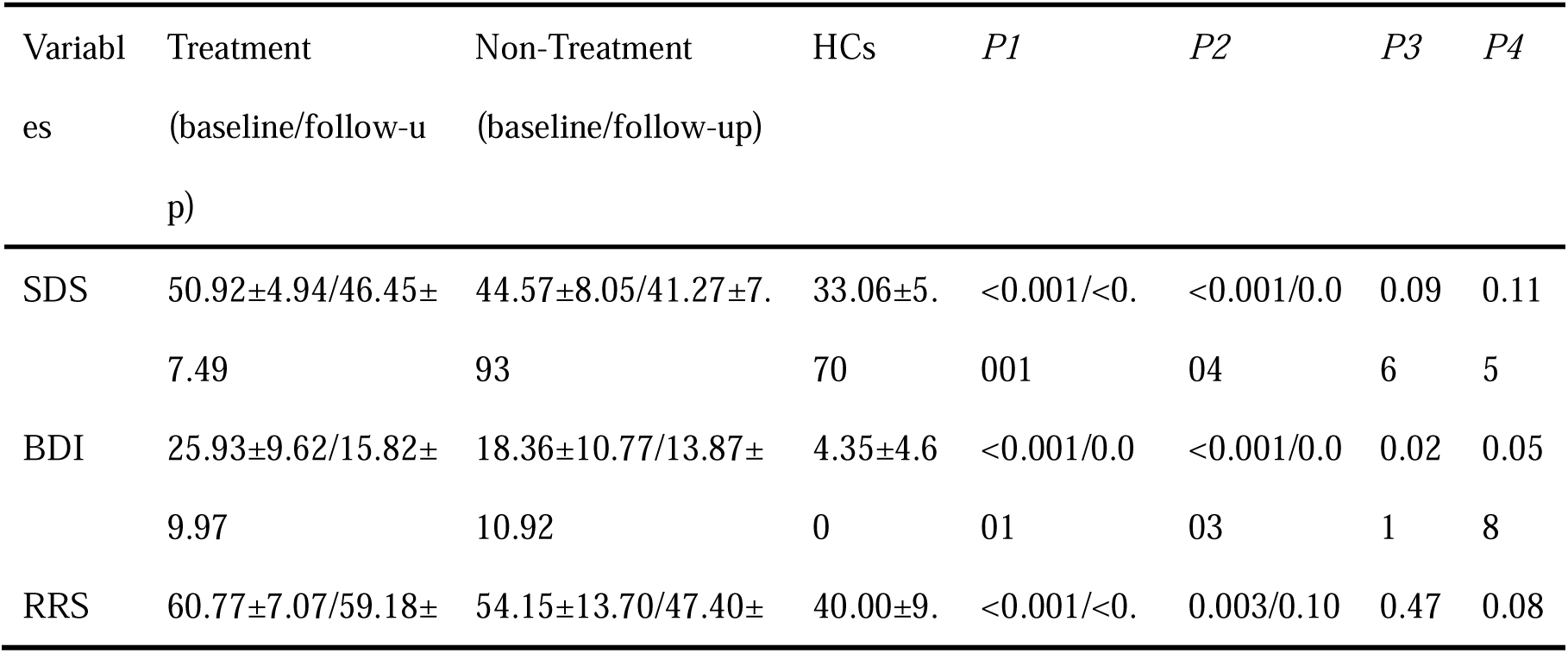
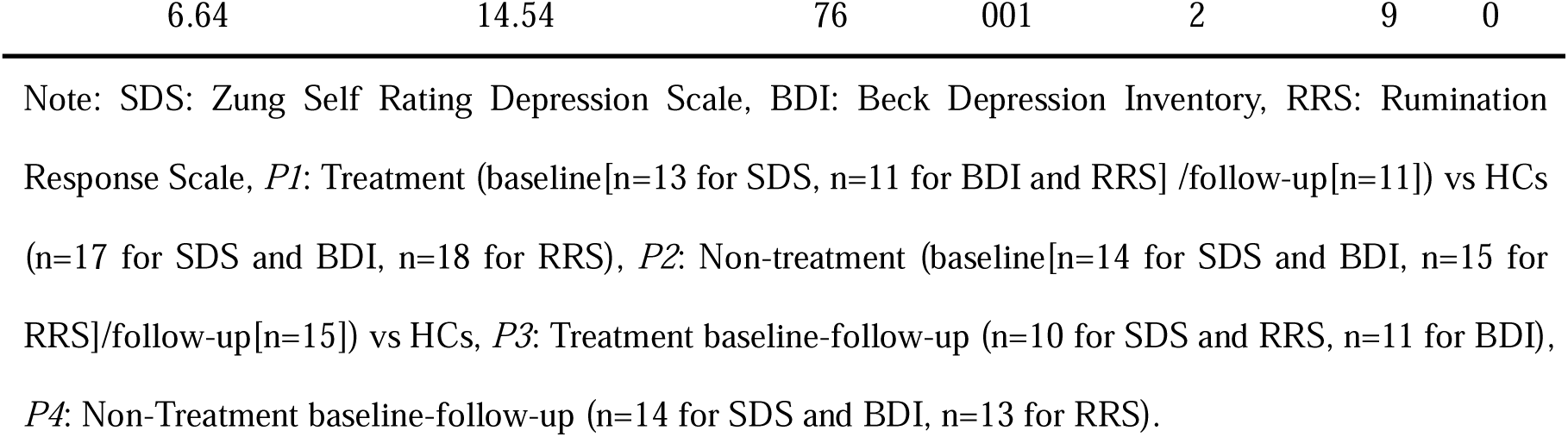
Clinical measures analysis.

### 3.2 BEN analysis

Compared to the HCs, DEP showed higher BEN in left DMPFC, left AMY/Putamen, and right hippocampus (HPC)/parahippocampal cortex (PHPC) (Fig. 1A). Functional decoding from a large-scale meta-analysis database (Neurosynth) for unthresholded two-sample *t*-test statistical map suggested positive association with emotion-related functional terms such as “face expression,” “fearful,” etc., and negative association with reward-related functional terms, such as “reward,” “value,” etc (Fig. 1B). Mean BEN values were extracted from each significant cluster, including the left DMPFC, left AMY/Putamen, and right HPC/PHPC for all participants. As illustrated in Fig. 1C, both Treatment and Non-Treatment demonstrated significantly higher baseline BEN compared to HCs in the left DMPFC (Treatment vs HCs: *t* = 4.96, *p* < 0.001, Cohen’s *d* = 1.727; Non-Treatment vs HCs: *t* = 2.93, *p* = 0.006, Cohen’s *d* = 0.999), left AMY/Putamen (Treatment vs HCs: *t* = 5.04, *p* < 0.001, Cohen’s *d* = 1.755; Non-Treatment vs HCs: *t* = 4.18, *p* < 0.001, Cohen’s *d* = 1.426), and right HPC/PHPC (Treatment vs HCs: *t* = 3.47, *p* = 0.002, Cohen’s *d* = 1.209; Non-Treatment vs HCs: *t* = 3.50, *p* = 0.001, Cohen’s *d* = 1.196). Additionally, Treatment exhibited significantly higher baseline BEN compared to Non-Treatment in the left DMPFC (*t* = 2.10, *p* = 0.045, Cohen’s *d* = 0.781). Repeated measures ANOVA revealed significant treatment-by-time interactions in all three ROIs (Fig. 1D, left DMPFC: *F* = 10.70, *p* = 0.003, left AMY/Putamen: *F* = 5.19, *p* = 0.031, right HPC/PHPC: *F* = 5.54, *p* = 0.026). Post hoc simple effects analysis demonstrated that Treatment showed significant BEN reduction in all three regions following intervention (Fig. 2A, left DMPFC: *t* = 4.94, *p* < 0.001, Cohen’s *d* = 1.784, left AMY/Putamen: *t* = 2.40, *p* = 0.032, Cohen’s *d* = 0.998, right HPC/PHPC: *t* = 3.82, *p* = 0.002, Cohen’s *d* = 0.609) and Non-Treatment exhibited no significant BEN changes at follow-up (Fig. 2B, left DMPFC: *t* = −0.52, *p* = 0.609, Cohen’s *d* = 0.178, left AMY/Putamen: *t* = 0.69, *p* = 0.504, Cohen’s *d* = 0.276, right HPC/PHPC: *t* = 0.27, *p* = 0.794,, Cohen’s *d* = 0.068). When comparing BEN at follow-up between patients and HCs, we found that the Non-Treatment continued to show significantly higher BEN than HCs in left DMPFC (*t* = 2.44, *p* = 0.020, Cohen’s *d* = 0.832), left AMY/Putamen (*t* = 4.19, *p* < 0.001, Cohen’s *d* = 1.432), and right HPC/PHPC (*t* = 3.31, *p* = 0.002, Cohen’s *d* = 1.130). In contrast, Treatment showed no significant differences in BEN compared to HCs in left DMPFC (*t* = 0.12, *p* = 0.905, Cohen’s *d* = 0.042) and left AMY/Putamen (*t* = 1.05, *p* = 0.303, Cohen’s *d* = 0.365) following treatment, while BEN in right HPC/PHPC (*t* = 2.43, *p* = 0.021, Cohen’s *d* = 0.848) remained significantly elevated compared to HCs. Furthermore, at follow-up, Non-Treatment demonstrated significantly higher BEN than the Treatment in both left DMPFC (*t* = 2.27, *p* = 0.032, Cohen’s *d* = 0.842) and left AMY/Putamen (*t* = 2.28, *p* = 0.031, Cohen’s *d* = 0.851).

**Figure 1.**
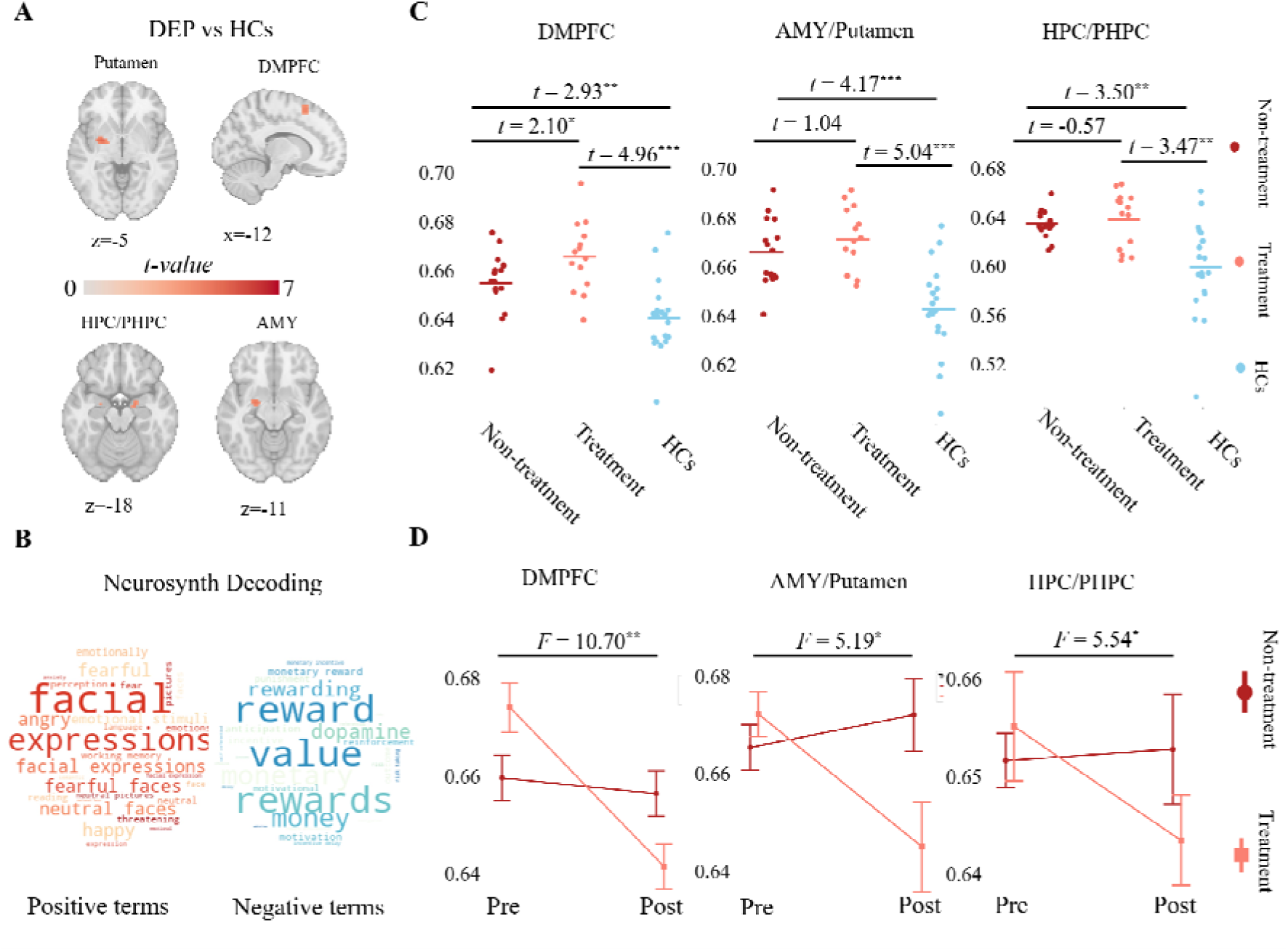
Increased resting BEN in mild to moderate depression was decreased by nonpharmacological treatment. **A.** Voxel-wise BEN differences between DEP and HCs at baseline. Red indicates higher BEN in DEP. **B.** Neurosynth decoding of the unthresholded *t*-map for DEP vs. HCs. Terms positively correlated with the pattern are shown in red, while negatively correlated terms appear in blue. ont size reflects correlation strength. **C.** Mean BEN values extracted from the left DMPFC, left AMY/Putamen, and right HPC/PHPC for each participant. Dark red indicates Non-Treatment, light red indicates the Treatment, and blue represents HCs; horizontal lines indicate group mean BEN. **D.** Treatment-by-time interaction plots of mean BEN in the left DMPFC, left AMY/Putamen, and right HPC/PHPC. Dark red and light red lines denote the Treatment and Non-Treatment, respectively; error bars represent standard error. Significance levels: \**p* < 0.05, \*\**p* < 0.01, \*\*\**p* < 0.001.

**Figure 2.**
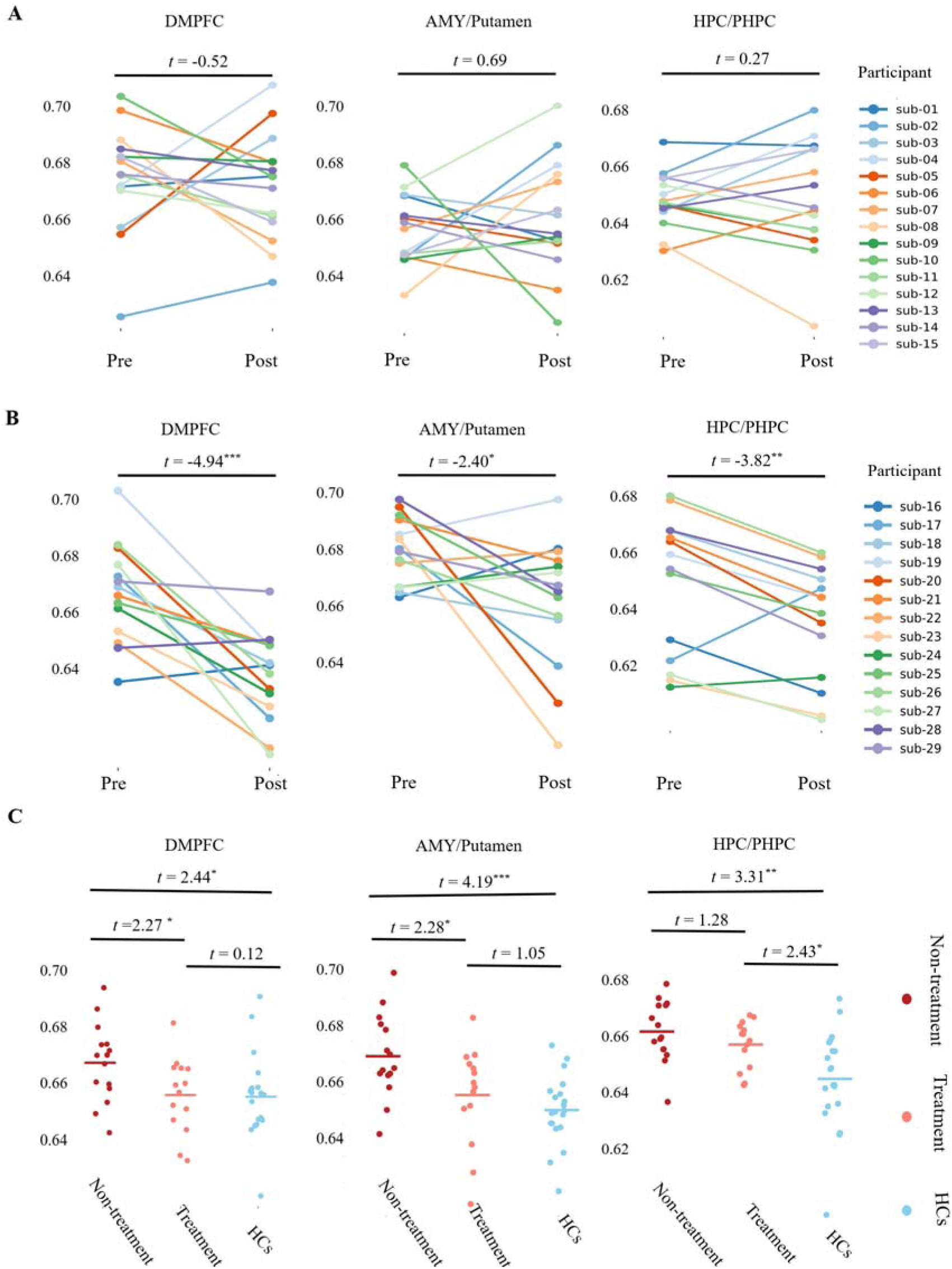
Longitudinal changes in BEN for each individual and comparison of BEN at follow-up between patients and HCs. **A.** Longitudinal changes in mean BEN (pre vs. post) within the Non-Treatment for the three regions of interest. Individual participant trajectories are shown. **B.** Longitudinal changes in mean BEN (pre vs. post) within the Treatment for the three regions of interest. Individual participant trajectories are shown. **C.** Differences of BEN at follow-up between patients and HCs. Dark red indicates the non-treatment group, light red indicates the treatment group, and blue represents HCs. Significance levels: \**p* < 0.05, \*\**p* < 0.01, \*\*\**p* < 0.001.

## 4. Discussion

This study demonstrates that alterations in BEN observed in individuals with mild to moderate depression can be reversed through nonpharmacological treatments. Specifically, increased BEN in the left DMPFC, left AMY/Putamen, and right HPC/PHPC, identified in patients with mild to moderate depression compared to HCs, was alleviated following nonpharmacological intervention at the individual level. Functional decoding based on Neurosynth analyses indicated that elevated BEN in these regions is primarily associated with emotion regulation and reward processing. The fronto-limbic and fronto-striatal networks, which are crucial for the top-down and bottom-up regulation of these processes, appear to be disrupted in depression. The observed increase in BEN within these networks may reflect dysfunction in emotion regulation and reward systems, which are core features of depressive symptomatology.

DMPFC and AMY play a crucial role in top-down regulation of emotions, serving as a key components in the emotional regulation circuitry (Berboth & Morawetz, 2021; Ma, Abelson, Okada, Taylor, & Liberzon, 2017; Morawetz, Hemetsberger, Laird, & Kohn, 2024; Pessoa, 2017). Lower BEN reflects more ordered and coherent neural activity, which may subsequently enhance the functional connectivity (FC) between the DMPFC and AMY, facilitating top-down emotional regulation. This perspective is supported by several lines of evidence. Firstly, the primary objective of NFB training is to enhance patients’ ability to regulate DMPFC. Secondly, greater activation in AMY during emotion processing tasks from task fMRI is associated with lower resting-state BEN in healthy adults (L. Lin et al., 2022). The regional BEN is negatively correlated with mean FC strength from 400 parcels of whole brain (D. Song, 2024), and in our recent study, we observed a negative correlation between BEN and FC within the left dorsolateral prefrontal cortex (DLPFC) (D.-H. Song & Wang, 2024b; D. Song & Wang, 2025). In another study, although we did not directly examine the relationship between BEN and FC, we found that subthreshold intermittent theta-burst stimulation (iTBS) applied to the left DLPFC resulted in lower BEN in the striatum (P. Liu et al., 2025) while another study revealed stronger FC between the left DLPFC and the striatum using the same dataset (Alkhasli, Sakreida, Mottaghy, & Binkofski, 2019). Thirdly, in an exploratory analysis using seed-based FC with the DMPFC as the region of interest (ROI), we found that, compared to HCs, depressed patients exhibited weaker FC between the DMPFC and AMY (uncorrected voxel-wise *p* < 0.05). However, after treatment, the FC between these regions increased (uncorrected voxel-wise *p* < 0.05). Compared to BEN, the changes in FC were less pronounced, possibly because these patients were in remission. More ordered neural activity (lower BEN) may facilitate increased interregional FC, with FC changes lagging BEN alterations. This suggests that BEN could serve as an early neural marker in the progression of depression and treatment. HPC/PHPC play a critical role in emotional memory processing and dynamically interact with the DMPFC and AMY(Girardeau, Inema, & Buzsáki, 2017; Jin & Maren, 2015; Qasim, Mohan, Stein, & Jacobs, 2023; Richardson, Strange, & Dolan, 2004). Reduced BEN in HPC/PHPC after nonpharmacological treatment is consistent with reduced BEN in HPC in MDD after an 8-week successful antidepressant treatment. Through the projections from AMY to HPC/PHPC, which subserves the creation and maintenance of emotional associations in memory, increased processing of negative emotion in AMY (Disner, Beevers, Haigh, & Beck, 2011) may ultimately lead to persistent negative associations (DeRubeis, Siegle, & Hollon, 2008; Dolcos, Iordan, & Dolcos, 2011; Sheline, Sanghavi, Mintun, & Gado, 1999). Reduced BEN in HPC/PHPC may facilitate the reconsolidation of emotionally disruptive memories. The observed attenuation of BEN in the putamen after treatment suggests improved reward processing efficiency. Notably, this effect may be mediated by the identified putamen-DMPFC reward circuitry, potentially reflecting optimized cortico-striatal communication.

Our results demonstrate that BEN can serve as an early biomarker for the onset, progression, and recovery of depression. Integrating our findings with clinical scale results, we observed that at baseline, Treatment exhibited significantly higher scores on depression clinical scales compared to Non-Treatment, with correspondingly higher BEN in left DMPFC, while no significant differences were found in left AMY/Putamen and right HPC/PHPC BEN between groups. At follow-up, however, depression clinical scale scores showed no significant differences between Treatment and Non-Treatment groups, whereas Treatment demonstrated significantly lower BEN in left DMPFC and left AMY/Putamen compared to Non-treatment, with levels comparable to HCs. In contrast, BEN of left HPC/PHPC showed no significant differences between Treatment and Non-Treatment, with both remaining elevated compared to HCs. These findings suggest that depression onset follows a bottom-up emotional dysregulation process, progressing from HPC/PHPC to AMY and finally to DMPFC, while recovery through nonpharmacological treatment follows a top-down cognitive control process, proceeding from DMPFC to AMY and then to HPC/PHPC (LeDuke, Borio, Miranda, & Tye, 2023). The lack of significant differences in depression clinical scale scores between Treatment andmay be attributed to the retrospective nature of clinical assessments, which typically reflect patients’ past emotional states, whereas treatment-induced changes in brain activity may not be immediately captured by these historical measures.

Our study also provides the possibility to compare the differences in neurological effects between pharmacological and nonpharmacological treatments. We have previously studied the relationship between BEN and pharmacotherapy in MDD (Liu, Song et al. 2020), and the results showed that relative to HCs, MDD had lower BEN in MOFC/sgACC, and lower MOFC/sgACC BEN was associated with less severe depression in MDD. This interesting relationship may be related to medication history. In addition, we also found that HPC is related to the improvement of medical treatment, possibly because medication can directly target the limbic system without relying on the top-down emotional regulation. In this study, CBT and NFB, two nonpharmacological treatments were used. A primary goal of CBT is to replace automatic emotional reactivity with more-controlled processing (Block, Klump et al. 2023), thus CBT might increase inhibitory executive control, helping to interrupt or dampen automatic limbic system reactions that consistent with neural mechanisms of the cognitive model of depression (Disner, Beevers et al. 2011). Some neuroimaging studies of changes in emotional reactivity during CBT are consistent with this formulation (Siegle, Carter et al. 2006, Siegle, Thompson et al. 2007, Ritchey, Dolcos et al. 2011). NFB is to promote the subject’s learning self-control over the activity in a selected brain area and, consequently, over the mental state corresponding with the activity in the given brain region (Linhartová, Látalová et al. 2019). Prefrontal cortex was selected as in the study, then this is reasonable to produce top-down emotion regulation. Nonpharmacological treatment might allow a resetting of DMPFC activity, to yield greater capacity for top-down emotion regulation when it is needed, medication might increase subcortical cingulate metabolism tonically, creating a bottom-up effect whereby relevant limbic regions are tonically inhibited.

Some limitations must be acknowledged. Our sample size was relatively small, only 14 participants received treatment, and due to the small post-separation sample sizes, we did not differentiate specific BEN changes between CBT and NFB treatments. Future research should focus on large sample sizes and specific neural circuits responsible for the effects of different treatment modalities. Nevertheless, we still observed increased BEN correlated with emotional terms using large scale decoding dataset. Furthermore, both cross-sectional and longitudinal analyses revealed consistent findings, demonstrating a significant reduction in BEN following nonpharmacological treatment in regions where BEN was relatively elevated compared to HCs. Notably, after nonpharmacological treatment, BEN in the DMPFC and AMY/Putamen of patients with mild to moderate depression was reduced to levels comparable to those of HCs, whereas no significant changes were observed in Non-Treatment. Additionally, we did not observe increased BEN in the MOFC/sgACC in this study, despite this area being widely recognized for aberrant activity in depression (Connolly et al., 2013; Drevets, Savitz, & Trimble, 2008; Fan, Wang, Wang, & Zang, 2024) and serving as a primary effective region for treatments for depression (Duprat et al., 2025; Morris et al., 2020; D. Song, Chang, Zhang, Peng, et al., 2019; D. Song et al., 2025). Unfortunately, significant signal dropouts occurred in the MOFC/sgACC in this dataset, which may represent the primary reason for our failure to detect BEN changes in this area.

In summary, patients with mild to moderate depression had increased BEN in DMPFC and limbic network, which was suppressed or reversed by nonpharmacological treatment. BEN in the DMPFC and limbic network could be a potential personalized marker for assessing onset, progression, and recovery processes of depression, and the effectiveness of nonpharmacological treatments.

## Data Availability

All data produced are available online at https://openneuro.org/datasets/ds002748 and https://openneuro.org/datasets/ds003007

https://openneuro.org/datasets/ds003007

## Data and code availability

The raw data is available from OpenNeuro coded as ds002748 (https://openneuro.org/datasets/ds002748) and ds003007 (https://openneuro.org/datasets/ds003007).

The BEN maps, FC maps and statistical maps are available at https://osf.io/ckagx/?view_only=1661268d1efe417a81a7b893904691da.

FSL is available at https://fsl.fmrib.ox.ac.uk/fsl/docs/#/. Nilearn is available at https://nilearn.github.io/dev/index.html.

SPM is available at https://www.fil.ion.ucl.ac.uk/spm/software/spm12/. BENtbx is available at https://www.cfn.upenn.edu/zewang/BENtbx.php.

Further updates related to the study will be available at https://github.com/donghui1119/BEN_mild_depression.

## Acknowledgements

We thank Dmitry D Bezmaternykh et al (D. D. Bezmaternykh et al., 2021) for releasing their dataset.

## Disclosures

The authors report no biomedical financial interests or potential conflicts of interest.

## CRediT authorship contribution statement

Donghui Song: conceptualization, data curation, data analysis, visualization, manuscript drafting and editing, project administration. Panshi Liu: visualization, validation, manu script editing. Xiaoye Ma: manuscript editing. Da Chang: manuscript editing. Ze Wang: conceptualization, manuscript editing, supervision.

## Notes

### Competing Interest Statement

The authors have declared no competing interest.

### Funding Statement

This study did not receive any funding

### Author Declarations

The study used (or will use) ONLY openly available human data that were originally located at https://openneuro.org/datasets/ds002748 and https://openneuro.org/datasets/ds003007

### Summary of Updates

In this updated version, we have reanalyzed the data and reviewed relevant research literature. Key improvements include the addition of an untreated control group in the longitudinal study of non-pharmacological treatments for depression. The results show that brain regions which exhibited differences between depressed patients and healthy controls were restored following treatment, whereas no significant changes were observed in untreated depressed patients. These new findings enhance the reliability of our conclusions. Additionally, we have systematically re-evaluated the literature to redefine the brain regions associated with entropy differences.

